# Promotion and Validation of Flipped Teaching Based on Video Conference in Standardized Training for Internal Medicine Residents

**DOI:** 10.1101/2022.07.20.22277876

**Authors:** Xiao-Yu Zhang

**Affiliations:** Section of Education, Shanghai Public Health Clinical Center, Fudan University, Shanghai, 201508, P. R. China; Department of Liver Disease, Shanghai Public Health Clinical Center, Fudan University, Shanghai, 201508, P. R. China; Public Health Education Professional Committee, Shanghai Preventive Medicine Association, Shanghai, P.R. China

**Keywords:** Flipped teaching, Internal medicine residents, Standardized training, Medical teaching

## Abstract

**Objective:** I aimed to verify the training effect of Flipped Teaching with Video Conference as carrier in the training of infectious diseases, and promote the application of this teaching mode in the standardized training for resident physicians.

**Methods:** The vertical management mode was adopted to carry out Flipped Teaching in the training, and the training of infectious diseases was carried out according to the requirements of the standardized training program for internal medicine resident. The residents trained in April were included in the trial group, and those trained in June were included in the validation group. The evaluation indexes of both groups were compared and analyzed.

**Results:** A total of 43 resident physicians participated in the training of infectious diseases by the Flipped Teaching based on Video Conference, all from tertiary hospitals. There were 19 participants in the trial group and 24 participants in the validation group, who carried out Flip Teaching with Video Conference as carrier for 31 times and 24 times respectively. The implementation rate of teaching plan was above 91% in both groups, and the attendance rate was above 98%; There was no significant difference on the attendance rate between the both groups, P>0.05. 952 effective feedback questionnaires were collected, including 395 in the trial group and 557 in the validation group. In terms of teaching quality, the indicators of “Rigorous teaching attitude” and “Punctual class” in the validation group were lower than those in the trial group, P < 0.05; And the other indicatiors were no significant difference, P>0.05; All feedback of the teaching quality indexes “good and very good” accounted for more than 96%. In the overall evaluation, there was no significant difference between the both groups on the index of “Suggestions for improvement”, P>0.05; And the “Praise highlight” of the validation group was higher than that of the trial group, P<0.05.

**Conclusion:** The Flipped Teaching based on Video Conference is generally effective for internal medicine residents to carry out training of infectious diseases, with good feedback and strong feasibility. This teaching model can be further promoted and applied in the standardized training for resident physicians.

## Introduction

In view of the fact that some standardized training bases for internal medicine residents did not meet the training standards of infectious diseases, those residents in the bases were assigned to a designated hospital to carry out the training of infectious diseases.[1] During the actual operation of the project, due to various subjective and objective reasons, the training cycle of infectious diseases training for internal medicine residents might be insufficient in a certain period; Therefore, I explored to apply a new training method as a supplement to the standardized training for internal medicine residents.

From April 1 to April 4, 2022, I began to explore Flipped Teaching with Video Conference as the carrier to carry out infectious disease training for internal medicine residents. This teaching mode was generally effective and could be used as a supplementary training method for standardized training of internal medicine residents, to make up for the lack of clinical training time for the residents due to holidays or emergencies.[2] Therefore, I conducted the infectious disease training for internal medicine residents with Flipped Teaching based on Video Conference in whole April.

In order to further verify the effect of Flipped Teaching with Video Conference as the carrier in the training of infectious diseases, I once again carried out the Flipped Teaching based on Video Conference for residents whose the original plan was arranged to the designated hospitals for infectious disease training in June 2022, which provided data support and experience summary for whether promoting the application of the teaching mode in the field of standardized training for resident physicians.

## Objects and Methods

### Subjects and Grouping

The study objects were internal medicine residents who planned to take training in infectious diseases training in the designated hospital in April and June. And they were also residents who had been participating in the standardized training in Shanghai. Their dispatched hospital had signed an Agreement on Joint Training of Internal Medicine Residents with the designated hospital.

Those residents who participated in the infectious diseases training with Flipped Teaching based on Video Conference in April and June were all included in the study, which was divided into two groups. Trial group: residents who participated in the infectious diseases training with Flipped Teaching based on Video Conference in April; Validation group: residents who participated in the infectious diseases training with Flipped Teaching based on Video Conference in June.

The informed consents of participants in internal medicine residents were obtained, including their data being used for the training and the research, and that this study was conducted in accordance with the Declaration of Helsinki.

### Construction of Flipping Teaching with Video Conference as the Carrier[2]

The Flipped Teaching with Video Conference as the carrier adopted “vertical management mode” for management[2,3], and training plan was formulated; All residents participated in teaching and evaluation. According to the training requirements of the Standardized Training Content and Standard for Resident Physicians (2021 Edition) -- Internal Medicine Training Rules, residents chose training topics and contents, and determined the teaching time within the training cycle; Teaching programme and implementation plans for the cycle would be identified after all training content and teaching time were determined; Residents developed PPT and clinical teaching based on the latest clinical guidelines and expert consensus. The whole teaching activity was carried out online by Video Conference, involving teaching organization, training, after-class discussion, teaching feedback, quality control and teaching management, etc.

### Evaluation Indexes and Criteria

This teaching model was validated from four aspects, including the implementation of the teaching plan, the attendance of the Flipped Teaching, the evaluation of teaching quality, and the overall evaluation of teaching.

Seven teaching plan indicators (topic selection based on training syllabus, teaching on the planned time, teaching on the planned content, making PPT fully, providing references, unifying the teaching content and training program, and participating in after-class discussion) were established to evaluate the implementation of the teaching plan. Two attendance indicators (online on time, and end on time) were established to evaluate the online attendance of the Flipped Teaching were completed. Nine teaching quality indicators (rigorous teaching attitude, punctual class, detailed and accurate teaching content, reasonable structure and clear process, highlighting teaching key points, clear teaching difficulties, accurate and refined language, combining theory with clinical practice, improving ability to analyze and deal with the disease) were established to evaluate the teaching quality. The overall evaluation of teaching adopted open questionnaire to evaluate each teaching without limit.

The teaching plan indicators and the attendance indicators were completed by the organizer, and the teaching quality indicators and the overall evaluation content were completed by every trainee for each teaching session. Among them, teaching plan indicators and teaching quality indicators were objective indicators, and they were derived from Teaching Evaluation Table of Standardized Residency Training; while overall evaluation indicators was subjective. The feedback of the over evaluation from open questionnaire was firstly classified according to the evaluation content. The details of classification as follow: If the content of a questionnaire feedback was pointing out deficiencies or needing improvement of one teaching session, this feedback was classified as “Improvement suggestions”; If all the content of a questionnaire feedback was praising highlights or learning achievements of one teaching session, this feedback was classified as “Praise highlights”; If the content of a questionnaire feedback was no special suggestions of one teaching session, this feedback was classified as “No special suggestions”.

### Software Application

Video Conference adopted Tencent Conference software to carry out online teaching, PPT playing, lecturing by residents, participating in questions, online answering, online check-in and discussion, dynamic monitoring, whole-process management, etc.

The software Questionnaire Star was used to develop 9 teaching quality indicators and 3 overall indicators, and carry out questionnaire star survey after class.

SPSS software version 23.0 (SPSS Inc. Chicago, IL, USA) was used for statistical analysis of the data. The data conforming to normal distribution were expressed as mean ± standard deviation to reflect the distribution of the study indicators. The counting data was represented by example (%) to reflect the composition ratio of the study indicators. Pearson Chi-square test or Fisher’s exact probability method was used for the counting data, and T test was used for data conforming to normal distribution. A P value of two-sided less than 0.05 was considered as statistically significant.

## Results

### Basic Information of Residents Participating in the Study

In this teaching research, a total of 43 residents participated in the infectious diseases training with Flipped Teaching based on Video Conference, all from tertiary hospitals. There were 19 resident physicians in the trial group, with an average age of 29.5 years, and male accounted for 47.4%; There were 24 residents in the validation group, with an average age of 28.5 years and 25.0% male. There were no significant differences in sexuality, age, education background, qualified as a licensed physician, and training phase between the both groups (P > 0.05). The detailed information is shown in *Table 1*.

**Table 1.** Baseline of Internal Medicine Residents Participating in Flipped Teaching

| Indicators | Trial group(n=19) | Validation group(n=24) | *p* value |
| --- | --- | --- | --- |
| Sexuality [male,n, (%)] | 9 (47.4) | 6 (25.0) | 0.126 |
| Age [years,Mean±standard deviation] | 29.5±3.1 | 28.5±3.0 | 0.287 |
| Education background |  |  |  |
| Bachelor degree[n, (%)] | 5 (26.3) | 11 (45.8) | 0.189 |
| Master degree[n, (%)] | 4 (21.1) | 4 (16.7) | 1.000 |
| Doctor degree[n, (%)] | 10 (52.6) | 9 (37.5) | 0.321 |
| Qualified as a licensed physician [n, (%)] | 17 (89.5) | 23 (95.8) | 0.575 |
| Training phase |  |  |  |
| First year of training[n, (%)] | 2 (10.5) | 0 (0) | 0.189 |
| Second year of training[n, (%)] | 12 (63.2) | 17 (70.8) | 0.594 |
| Third year of training[n, (%)] | 5 (26.3) | 7 (29.2) | 0.836 |
| From tertiary hospitals [n, (%)] | 19 (100) | 24 (100) | – |

### Evaluation of Teaching Plan Implementation

The trial group carried out 31 Flipped Teaching sessions, and the verification group carried out 24 Flipped Teaching sessions. In the trial group and the validation group, the times of “teaching on the planned time” were 29 times and 22 times, accounting for 93.5% and 91.7%, respectively; and there was no significant difference between the both groups in other teaching performance indicators, including”topic selection based on training syllabus”, “teaching on the planned content”, “making PPT fully”, “providing references”, “providing references”, and “Providing references”, P > 0.05. In the both groups, the proportion of the other teaching performance indicators conforming to the training plan was 100%. The detailed information is shown in *Table 2*.

**Table 2.** Statistical Table of Evaluation of Teaching Plan Implementation

| Teaching plan indicators[n, (%)] | Trial group (n=31) | Validation group (n=24) | *p* value |
| --- | --- | --- | --- |
|  | Coincidence times (%) | Coincidence times (%) |  |
| Topic selection based on training syllabus | 31 (100) | 24 (100) | – |
| Teaching on the planned time | 29 (93.5) | 22 (91.7) | 1.000 |
| Teaching on the planned content | 31 (100) | 24 (100) | – |
| Making PPT fully | 31 (100) | 24 (100) | – |
| Providing references | 31 (100) | 24 (100) | – |
| Unifying the teaching content and training program | 31 (100) | 24 (100) | – |
| Participating in after-class discussion | 31 (100) | 24 (100) | – |

### Teaching Attendance

The attendance was checked for the whole process of the Flipped Teaching based on Video Conference, while “online on time” and “end on time” two time nodes were included in the statistics. In the trial group, the total attendance times of “online on time” and “end on time” were 585 times and 588 times, accounting for 99.3% and 99.8% respectively. In the validation group, the total attendance times of “online on time” and “end on time” were 566 times and 574 times, accounting for 98.3% and 99.7% respectively. There was no significant difference in attendance between the both group at the above two time nodes, P > 0.05. The detailed attendance is shown in *Table 3*.

**Table 3.** Total Attendance of of Flipped Teaching

| Attendance indicators[n, (%)] | Trial group (n=589) | Validation group (n=576) | *p* value |
| --- | --- | --- | --- |
|  | Total attendance times(%) | Total attendance times(%) |  |
| Online on time | 585 (99.3) | 566 (98.3) | 0.098 |
| End on time | 588 (99.8) | 574 (99.7) | 0.621 |

### Evaluation of Teaching Quality

A total of 952 effective feedback questionnaires were collected, including 395 in the trial group and 557 in the validation group. The feedback of “good and very good” on the index of “rigorous teaching attitude” in the validation group was lower than that in the trial group, accounting for 96.9% and 99.7% respectively, P=0.002. In terms of “punctual class” index, the feedback of “good and very good” in the validation group was lower than that in the trial group, accounting for 97.8% and 99.5% respectively, P=0.037. There was no significant difference in attendance between the both group on the seven indexes of “detailed and accurate teaching content”, “reasonable structure and clear process”, “highlighting teaching key points”, “clear teaching difficulties”, “accurate and refined language”, “combining theory with clinical practice”, and “Improving ability to analyze and deal with the disease”, P > 0.05. In general, all the indicators of the teaching quality evaluated as “good and very good” accounted for more than 96%. The detailed evaluation of teaching quality is shown in *Table 4*.

**Table 4.**
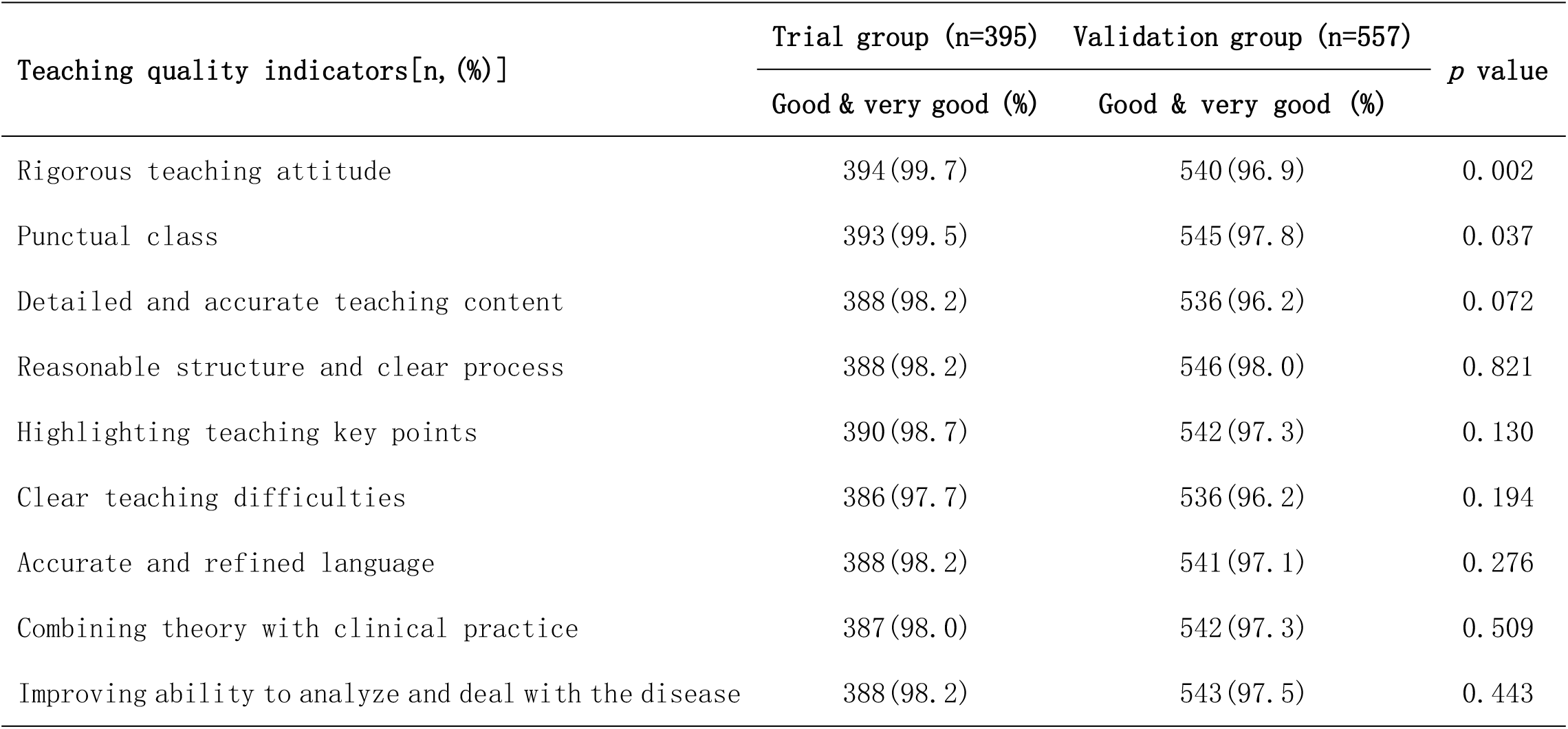
Statistical Table of Flipped Teaching Quality Evaluation

### Overall Evaluation of Flipped Teaching

In the overall evaluation of Flipped Teaching based on Video Conference, a total of 952 valid questionnaires were collected through the questionnaire star. 25 and 49 questionnaires with feedback of “improvement suggestions” were collected in the trial group and the validation group, accounting for 6.3% and 8.8% respectively; And there was no significant difference between the both groups, P > 0.05. 144 and 120 questionnaires with feedback of “none” were collected in the trial group and the validation group, accounting for 36.5% and 21.5%, respectively; And the feedback of “none” from the validation group was lower than those from the trial group, P < 0.001. 226 and 388 questionnaires with feedback of “Praise highlights” were collected in the trial group and the validation group, accounting for 57.2% and 69.7%, respectively; And the feedback of “Praise highlights” from the validation group was higher than those from the trial group, P < 0.001. The detailed overall evaluation of feedback is shown in *Table 5*.

**Table 5.** Overall Evaluation of Flipped Teaching

| Evaluation indicators[n, (%)] | Trial group (n=395) | Validation group (n=557) | *p* value |
| --- | --- | --- | --- |
|  | Good & very good (%) | Good & very good (%) |  |
| Improvement suggestions | 25(6.3) | 49(8.8) | 0.161 |
| None | 144(36.5) | 120(21.5) | <0.001 |
| Praise highlights | 226(57.2) | 388(69.7) | <0.001 |

## Discussion

This research on Flipped Teaching with Video Conference as the carrier was based on the good results achieved by the teaching model in the early stage[2]. The implementation and evaluation of the Flipped Teaching based on Video Conference in April and June were compared and analyzed to further verify the training effect and repeatability of this teaching model. To this end, we carried on the following six aspects of analysis and elaboration to this study.

### Highlights of the Study

The infectious diseases training was a necessary part of the standardized training for internal medicine residents. For internal medicine residents who could not carry out the standardized training of infectious diseases according to the original plan, the Flipped Teaching based on Video Conference was carried out to effectively make up for the shortage of the actual training period in the infectious diseases. Through scientific design and teaching practice, the training effect and repeatability of this model were verified. This teaching model could be conducted to the smooth progress of standardized training for residents in the region and had obvious whole concept and social significance.

### Key Points of the Study

The teaching research adopted “vertical management mode”. Since the “vertical management mode” established had gained good experience and evaluation in the field of standardized training for public health physicians and internal medicine residents, the organizer of which should have managerial, clinical and research capabilities[2-3]. Therefore, the application of this management mode in this teaching was an important basis to ensure the smooth development of this teaching research and obtain good feedback. As the residents participating in the training of infectious diseases came from many hospitals, the supports from the teaching administrative departments and training bases of all dispatched hospitals were also important factors to ensure the smooth progress of the teaching. Furthermore, according to the requirements of the training syllabus and the training purpose, it was also the key to clarify the training contents and references to ensure the quality of Flipped Teaching based on Video Conference.

### Comparison with Related Literature

With the publication of the paper “Educators propose ‘flipping’ medical training” in 2012[4], Flipped Teaching had been studied and applied more and more in the field of medical education, such as Cardiology[5], Internal Medicine[6,7], Anaesthesiology[8], Rheumatology[9], Pathology[10], Radiology[11], Nutriology[12], dermatologic surgery[13,14], Nursing[15], et cetera, and teaching and research achievements of different degrees were achieved. Generally, in the evaluation of Flipped Teaching from the above literature cited, more initiators and researchers were more likely to draw positive conclusions from impressive teaching achievements and put forward the positive significance of Flipped Teaching.[5-15] However, there were different opinions on Flipped Teaching for that some participants themselves were not familiar with the organization and implementation of Flipped Teaching, some residents lacked clinical knowledge reserve, and the preparation time of flipped teaching was relatively long[16.17].

In view of the fact that there was no unified definition and implementation of Flipped Teaching in the teaching field, and Inconsistent background and application domain of this kind of teaching, there were great differences in teaching practice. Therefore, I had made a detailed definition and practice of this Flipped Teaching; Including the following aspects: “vertical management mode” was adopted for this teaching, clinical teaching was carried by Video Conference, the teaching subjects were led by residents, the teaching contents were based on the requirements of the Standardized Training Content and Standard for Resident Physicians, the teaching reference materials were based on the latest clinical guidelines and experts consensus of infectious diseases, characteristic teaching programs and training plans were developed by all participants, every Flipped Teaching was managed in the whole process, all joint training unit of those residents needed to participate in the teaching management and assessment of residents, and the effect of each Flipped Teaching would be evaluated by all residents and the organizer.

### Interpretation of Relevant Data in the Study

There was no significant difference in baseline data between the both groups (P > 0.05), indicating that the study was comparable and the conclusions were reliable. The implementation of the teaching plan was good; In addition to “teaching on the planned time”, the completion rate of the other teaching plan indicators reached 100%; Some residents could not carry out teaching on time; which was also related to that some residents were unfamiliarity with teaching software, or they were performing part of the clinical work at that point. The attendance rates of the both groups was high, above 98%; And this was higher than Flipped Teaching in other areas, where attendance was 30-80%[4], which might be related to the cooperation of residents, the support of joint training units and the influence of the teaching organizers. The teaching quality evaluation was generally good, and the feedback of all indicators were “good and very good” accounted for more than 96%; Among them, the evaluation of “rigorous teaching attitude” and “punctual class” by residents in June was lower than that in April (P < 0.05), which might be related to the fact that most of the residents in June were also engaged in clinical work, while most of the internal medicine residents were on vacation in April. In terms of the overall evaluation of the teaching, there was no significant difference between the two groups in the evaluation of “suggestions for improvement” (P > 0.05); The evaluation of “praise highlights” put forward by residents in June was higher than that of internal medicine residents in April (P < 0.05), which reflected that the Flipped Teaching based on Video Conference was more recognized by later residents along with this teaching model continuing to advance.

### Advantages of the Study

The rapid increase in medical knowledge and clinical workload had resulted in a relatively limited amount of time for clinicians and residents to undertake effective training. The Flipped Teaching based on Video Conference could make up for the defects. Teaching and training could be arranged during off-peak hours by this teaching model; That could not only get rid of the space limitation of clinical training, but also reduce the time constraint of clinical teaching. Therefore, the Flipped Teaching based on Video Conference had good application value.

In order to further verify the feasibility and effectiveness of the Flipped Teaching based on Video Conference, both groups of queues were designed to conduct control study. The infectious diseases training by the Flipped Teaching based on Video Conference for residents was carried out again in June, and compared with the same teaching method for residents in April. The repeatability of the training effect of Flipped Teaching based on Video Conference was further verified. All the above embodied the scientific nature of this teaching research.

### Limitations of the Study

The Flipped Teaching based on Video Conference adopted online whole-process monitoring, and three time nodes of “online on time”, “middle roll call” and “end on time” were selected to be included in the statistics. However, due to incomplete data of “middle roll call”, the data of “middle roll call” was not included in this study for attendance statistical analysis. This was the deficiency of this teaching research. Moreover, since the number of residents participating in the Flipped Teaching based on Video Conference was small, this model was suitable for the training of small groups. For a large number of resident training, a new study with larger sample would be needed to support.

At the beginning of this teaching model, the project had high requirements for the teaching organizer. The organizer owned management background and clinical work background, and also needed the education background and teaching background; At the same time, it called for good scientific research ability for teaching evaluation and promotion. The high standard requirements for the organizer at the beginning of the project were important foundations to ensure the smooth progress of this teaching mode. However, once the teaching had been successfully promoted, a systematic mode of Flipped Teaching was formed, and the follow-up work would not be so high demand for the organizers.

## Conclusions

It is generally feasible to carry out infectious disease training for internal medicine residents with Flipped Teaching based on Video Conference, and the teaching plan execution and teaching feedback are good, and the training effect is obvious. This teaching model can be further promoted and applied in the standardized training of residents.

## Data Availability

All data produced in the present study are available upon reasonable request to the authors

## Ethical Approval and Consent to Participate

Informed consents of participants in the standardized training for internal medicine residents were obtained for the training and the study. The study received Institutional Review Board (IRB) approval by the Shanghai Public Health Clinical Center Ethics Committee. The IRB number was No. 2021-S026-01.

## Acknowledgments

This study was supported by the internal medicine residents who had participated in the standardized training for residents in Shanghai. Thanks to the teaching administration department and the resident standardized training base of the united training hospital for their supports.

## Authors’ Contributions

Xiao-Yu Zhang made conception, design, acquisition of data, analysis and interpretation of data, drafted and revised the manuscript, and agreed to be accountable for all aspects of the work.

## Consent for publication

The author has read and agreed to the published version of the manuscript.

## Literature share statement

The study included in the manuscript submitted to the journal is transparent. Consent from the corresponding author is required for any institution or individual to reprint this document.

## References

[1] Shanghai health and family planning commission,Notice on strengthening standardized training of residents in Shanghai for rotation training of infection department of internal medicine residents. http://wsjkw.sh.gov.cn/kjjy2/20180815/0012-57913.html

[2] Zhang XY. Application and Evaluation of Flipped Teaching Based on Video Conference in Standardized Training for Internal Medicine Residents during Vacation. bioRxiv; 2022. doi: 10.1101/2022.04.17.488599

[3] Zhang XY. Rapid Clinical Promotion Model of Standardized Training for Public Health Physicians in China. Adv Med Educ Pract. 2021;12:463–471. doi:10.2147/AMEP.S306737

[4] Vogel L. Educators propose “flipping” medical training. CMAJ. 2012;184(12):E625–E626. doi:10.1503/cmaj.109-4212

[5] Narang A, Velagapudi P, Rajagopalan B, et al. A New Educational Framework to Improve Lifelong Learning for Cardiologists. J Am Coll Cardiol. 2018;71(4):454–462. doi:10.1016/j.jacc.2017.11.045

[6] Bonnes SL, Ratelle JT, Halvorsen AJ, et al. Flipping the Quality Improvement Classroom in Residency Education. Acad Med. 2017;92(1):101–107. doi:10.1097/ACM.0000000000001412

[7] Wittich CM, Agrawal A, Wang AT, et al. Flipped Classrooms in Graduate Medical Education: A National Survey of Residency Program Directors. Acad Med. 2018;93(3):471–477. doi:10.1097/ACM.0000000000001776

[8] Kurup V, Sendlewski G. The Feasibility of Incorporating a Flipped Classroom Model in an Anesthesia Residency Curriculum-Pilot Study. Yale J Biol Med. 2020;93(3):411-417. Published 2020 Aug 31.

[9] Wade SD, Freed JA, Kyttaris VC, Saunders S, Hausmann JS. Implementing a Virtual Flipped Classroom in a Rheumatology Fellowship Program [published online ahead of print, 2021 Sep 22]. Arthritis Care Res (Hoboken). 2021;10.1002/acr.24791. doi:10.1002/acr.24791

[10] Koch LK, Chang OH, Dintzis SM. Medical Education in Pathology: General Concepts and Strategies for Implementation. Arch Pathol Lab Med. 2021;145(9):1081–1088. doi:10.5858/arpa.2020-0463-RA

[11] Tan N, Bavadian N, Lyons P, Lochhead J, Alexander A. Flipped Classroom Approach to Teaching a Radiology Medical Student Clerkship. J Am Coll Radiol. 2018;15(12):1768–1770. doi:10.1016/j.jacr.2018.07.017

[12] Riddle E, Gier E, Williams K. Utility of the Flipped Classroom When Teaching Clinical Nutrition Material. J Acad Nutr Diet. 2020;120(3):351–358. doi:10.1016/j.jand.2019.09.015

[13] Tassavor M, Shah A, Hashim P, Torbeck R. Flipped classroom curriculum for dermatologic surgery during COVID-19: A prospective cohort study. J Am Acad Dermatol. 2021;85(5):e297–e298. doi:10.1016/j.jaad.2021.06.891

[14] Liu KJ, Tkachenko E, Waldman A, et al. A video-based, flipped classroom, simulation curriculum for dermatologic surgery: A prospective, multi-institution study. J Am Acad Dermatol. 2019;81(6):1271–1276. doi:10.1016/j.jaad.2019.03.078

[15] Hu R, Gao H, Ye Y, Ni Z, Jiang N, Jiang X. Effectiveness of flipped classrooms in Chinese baccalaureate nursing education: A meta-analysis of randomized controlled trials. Int J Nurs Stud. 2018;79:94–103. doi:10.1016/j.ijnurstu.2017.11.012

[16] Hew KF, Lo CK. Flipped classroom improves student learning in health professions education: a meta-analysis. BMC Med Educ. 2018;18(1):38. Published 2018 Mar 15. 413 doi:10.1186/s12909-018-1144-z

[17] Evans L, Vanden Bosch ML, Harrington S, Schoofs N, Coviak C. Flipping the classroom in health care higher education: a systematic review. Nurse Educ 2019;44:74–78. 417 doi:10.1097/NNE.0000000000000554

